# No stimulant? No Problem: Analyzing Black Market Sales of Atomoxetine on StreetRx

**DOI:** 10.1101/2022.12.19.22283638

**Authors:** Sophie A. Roe, Dayna S. DeSalve, Brian J. Piper

**Affiliations:** Geisinger Commonwealth School of Medicine

## Abstract

**Objective:** The purpose of this report was to analyze black market diversions of atomoxetine, a non-stimulant medication for ADHD, submitted to StreetRx.

**Method:** Data related to black market atomoxetine purchases entered on StreetRx between 2015 and 2019 were analyzed. Data included generic drug name, dosage, purchase price, and date and location in the United States. The mean price per milligram was determined and a heatmap generated.

**Results:** The average price per milligram cost of 113 diverted atomoxetine submissions was $1.35 (± $2.76 SD) (Median = $0.05, Min = $0.01, Max = $20.00). The states with the most submissions included Michigan (11), Pennsylvania (9), Indiana (8), and Ohio (8).

**Conclusion:** Furture qualitative studies should investigate reasons why individuals are motivated to purchase atomoxetine, a non-stimulant medication, on the black market (recreational vs. nootropic vs. other clinical uses).

## Introduction

Atomoxetine became the first non-stimulant medication to receive US Food and Drug Administration (FDA) approval for the treatment of attention deficit/hyperactivity disorder (ADHD) in 2002. It is typically used as a second-line medication for adults and, less commonly, children with contraindications to first-line stimulant medications (methylphenidate) (Cortese et al., 2018). Given that atomoxetine is unscheduled, indicating a low abuse potential, it may be particularly useful for pediatric patients and their families that would prefer to avoid a controlled substance (Garnock-Jones et al., 2009). When administered at approved dosages, atomoxetine may require several weeks of treatment before therapeutic effects are attained, in comparison to methylphenidate and other amphetamines, which have almost immediate effects (Fedder et al., 2022; Jaeschke et al., 2022). Atomoxetine carries an FDA warning of potential suicidal ideation (irritability, agitation, thoughts of suicide or self-harm) in children and adolescents. Therefore, children and adolescents taking the medication should be monitored for behavioral changes, suicidal thinking or behavior, and/or clinical worsening (Pliszka, 2007; Brown, 2018). If any of these rare adverse effects are identified, the patient should cease use immediately (Pliszka, 2007). These potential adverse effects combined with a lower efficacy reported in some studies have resulted in atomoxetine being considered second to stimulant medications for treating uncomplicated ADHD in some comparative studies (Fu et al, 2022). Nonetheless, atomoxetine may be preferred for the treatment of patients with comorbid anxiety and patients with a drug misuse history, both of which increase the risks associated with stimulant use (Garnock-Jones et al., 2009). It may also be preferred in patients that experience severe side effects to stimulant use, such as mood instability or tic disorders (Biederman et al., 2004; Pliszka, 2007).

Contrary to the first line stimulants, which are classified as Schedule II drugs in the US, atomoxetine is unscheduled based on its “low abuse potential” reported in several studies (Bymaster et al., 2022; Upadhyaya et al., 2013). The classification is primarily due to its mechanism of action as a selective inhibitor of the presynaptic norepinephrine transporter that has no notable affinity for the central receptors (dopamine transporters, GABA_A_ receptors, and opioid μ receptors) (Upadhyaya et al., 2013). Increasing dopamine produces the “high” associated with methylphenidate and lisdexamfetamine, which is not seen with atomoxetine use (Fedder et al., 2022; Upadhyaya, 2013). However, findings from several rat studies have determined that atomoxetine increases dopamine in the prefrontal cortex and may also exert its effect through non-dopaminergic mechanisms involving prefrontal histamine release (Horner et al., 2006).

Since 2010, StreetRx has served as a crowdsourced data platform through which users submit information on Black Market Drug exchanges (“About StreetRx,” n.d.). The large number of publications that have utilized crowdsourced opioid exchange data from StreetRx evidence its ability to provide valid, up-to-date street prices for diverted prescription drugs (Hswen, Y., Zhang, A., & Brownstein, J. S. 2020; Freifeld, C et al., 2013; Lebin et al., 2018). StreetRx gathers information on price, quantity, and location of a given diverted drug exchange directly from purchasers in the United States, Australia, Canada, France, Germany, Italy, Spain, and the United Kingdom. StreetRx is partnered with Researched Abuse, Diversion, and Addiction-Related Surveillance System (RADARS), which also collects data on prescription drugs that are abused, misused, and diverted (“About StreetRx,” n.d.). The objective of this study was to characterize the pattern of atomoxetine reports to StreetRx.

## Methods

### Procedures

Access to StreetRx data reports was granted via a data use agreement between Geisinger Commonwealth School of Medicine and Rocky Mountain Poison and Drug Safety Department of the Denver Health and Hospital Authority. Data included the generic drug name, drug dosage, purchase price, and date and location of submission in the United States from 2015 to 2019 for atomoxetine. Procedures were approved by the Geisinger Institutional Review Board.

### Analysis

The mean price per milligram was computed using GraphPad Prism v. 9.4.0. Figures were constructed using GraphPad Prism v. 9.4.0. Heatmapper was used to represent the location of submissions (Babicki et al., 2022).

## Results

A total of 113 atomoxetine submissions were made to StreetRx from 2015 to 2019. The average price per milligram of diverted atomoxetine (including generic and brand name) was $1.35 with a standard deviation of $2.76 (Figure 1, Supplemental Figure 1). Generic atomoxetine averaged a modest price per milligram of $0.08 (N=8; SD=$0.03) and brand name atomoxetine (Strattera) averaged a price per milligram of $1.45 (N=105; SD=$2.84). The most submissions occurred in Michigan (11) followed by Pennsylvania (9), Indiana (8), and Ohio (8). Thirteen states (AK, AR, DE, HI, IA, LA, MS, MT, NM, ND, RI, VT, WI) did not have any submissions (Figure 2).

**Figure 1.**
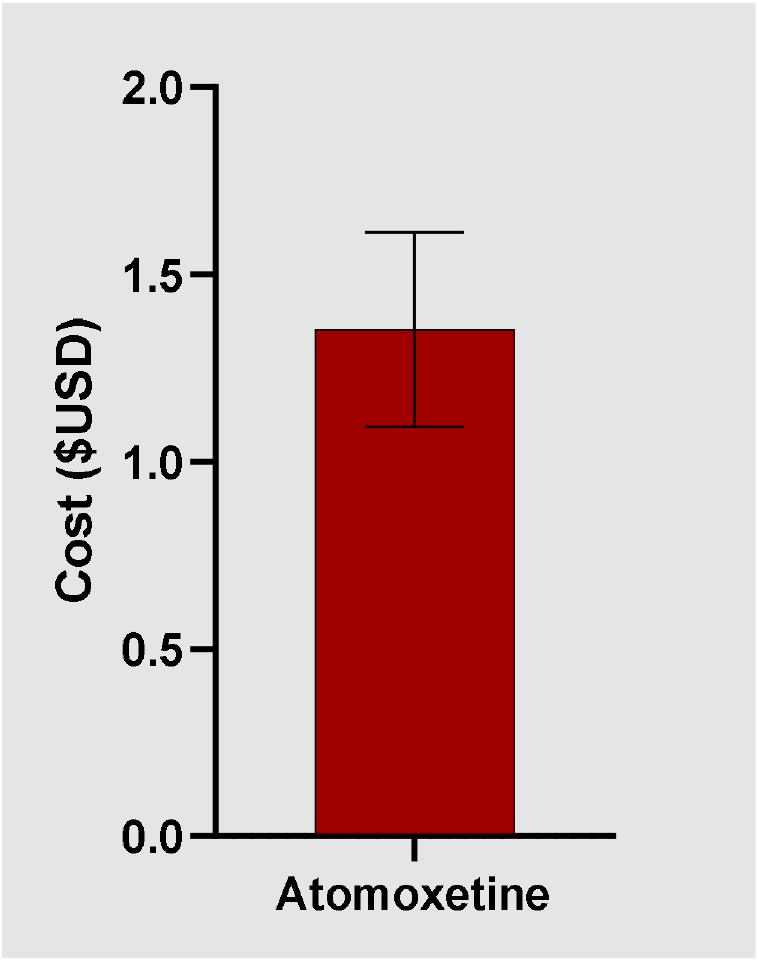
Average (±SEM) cost per mg of diverted atomoxetine as reported to StreetRx.

**Figure 2.**
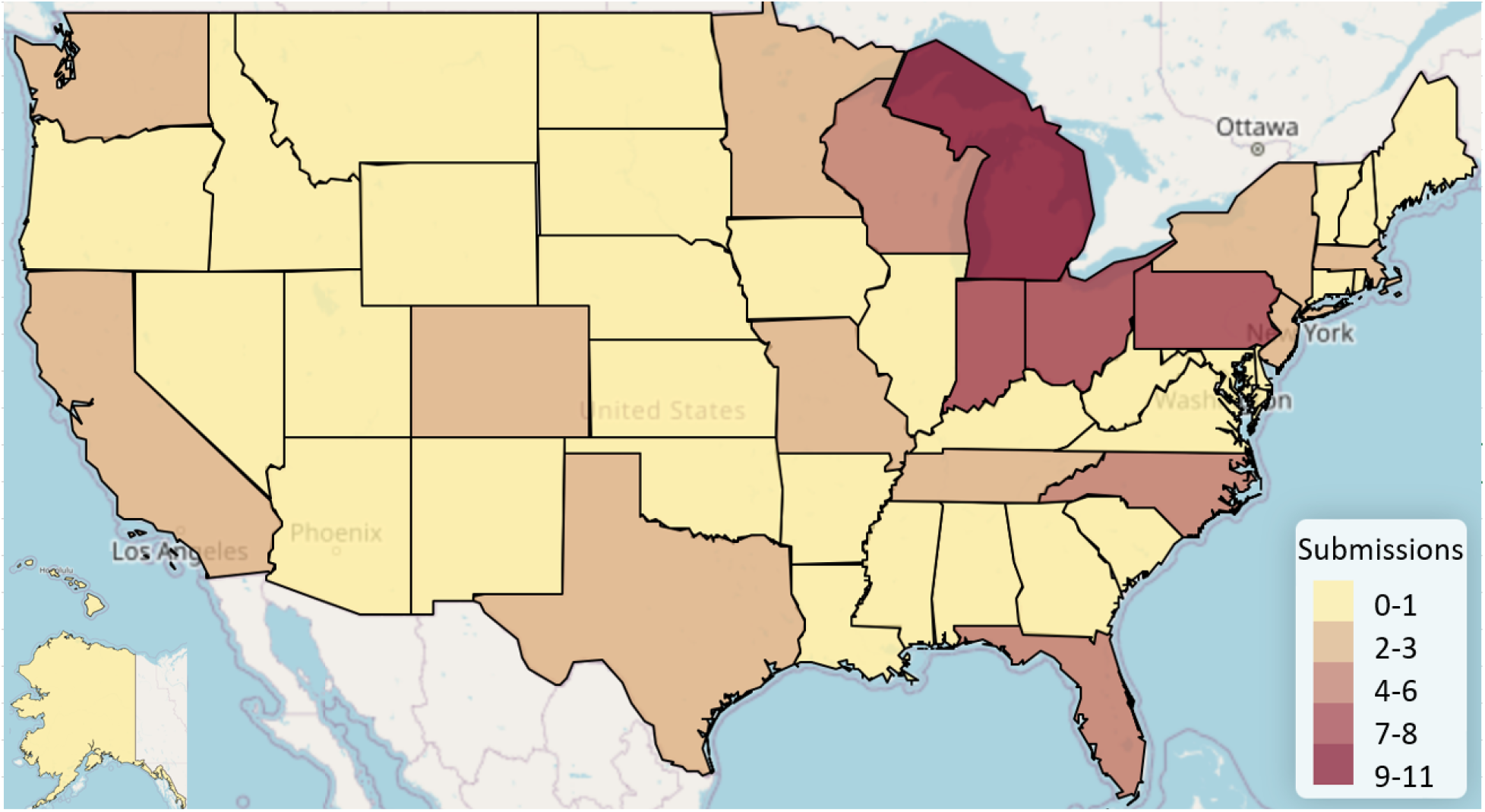
Heat map displaying diverted atomoxetine submissions per state submitted to StreetRx.

## Discussion

This novel report analyzed 113 black market atomoxetine exchanges, which mostly took place in midwestern states (Michigan, Ohio, Indiana, and Pennsylvania). Given that atomoxetine has not been shown to act through the striatal dopaminergic pathways through which drugs of abuse typically act, it may be suprising that people would choose to purchase it over centrally-acting agents on the black market (i.e. amphetamines). It is even more suprising that people would choose to purchase atomoxetine at an average price of $1.35/mg, rather than the centrally-acting stimulant drugs amphetamine or lisdexamfetamine, which averaged $0.53 and $0.28 per mg per exchange, respectively (Brasch et. al., in preparation). According to GoodRx, the lowest prescribed price per milligram (ppm) cost of atomoxetine is $0.024/mg, which is much lower than its black market price of $1.35/mg. Also noteworthy was that there were thirteen-fold more submissions for brand name than generic atomoxetine and the price per mg of generic atomoxetine was one-eighteenth that of the brand name. Therefore, it is evident that individuals who have a prescription and subsequently sell atomoxetine on the street could be making a substantial profit.

There are several possible reasons why there could be a demand for diverted atomoxetine. First, non-ADHD populations may desire cognitive enhancement but prefer not to experience certain effects of stimulants (Malík & Tlustoš, 2022). It is also possible that some individuals experience desirable neuropsychotropic effects even though atomoxetine is not a stimulant. There is an Erowid report of a teenager taking more than the recommended dose and becoming hyperverbal, an experience he rated favorably (GBVsoldier, 2008). Additional anonymous reports describe the synergistic effects of atomoxetine and caffeine or its synergistic effects with mirtazapine (Doctor Faust, 2009; Terra, 2014). Second, atomoxetine’s noradrenergic effects could be used as a party drug with the intention of offsetting the depressant effects of alcohol. Additionally, several studies have reported that CYP2D6 genotype can affect atomoxetine efficacy. Poor CYP2D6 metabolizers have been found to experience superior treatment outcomes as measured by reduction in ADHD symptoms, but also an increase in medication-related side effects relative to extensive CYP2D6 metabolizers (Brown et al., 2019; Michelson et al, 2007). It is possible that certain purchasers captured in this study were motivated by their superior response to atomoxetine based on their CYP2D6 genotype. As for possible drug interactions, atomoxetine is metablized by the P450 2D6 enzyme, thus any other medications (paroxetine, fluoxetine) that inhibit this enzyme can raise the serum atomoxetine levels by three or four-fold (Sawant & Daviss, 2004). It is possible that some StreetRx atomoxetine entries were by individuals who combined it with any of these drugs in order to produce an unusual or enhanced effect.

It is also possible that a subset of black market atomoxetine purchasers who were well-versed in the “off-label” uses of the drug purchased it to self-treat a number of conditions. For example, a number of studies have documented atomoxetine’s effectiveness for treating obesity and eating disorders (particularly binge eating disorder), as well as concurrent substance disorders and ADHD (Kollins, 2008; Cunnill, 2022). In the past, it was used to aid tobacco cessation and treat the symptoms of nicotine withdrawl (Ray et al., 2009). It is also possible that purchasers were using atomoxetine to combat the effects of alcohol, produce sexual effects, and limit body weight, given existing research documenting all of these uses (Gadde et al., 2006). Finally, it is possible that some purchasers are simply unaware of atomoxetine’s nonstimulant status and believe they are purchasing something no different from amphetamines.

The next logical question is: to what extent should self-treatment with atomoxetine via black market purchasing be of concern from a public safety perspective? While some atomoxetine purchases documented in this study may represent patients attempting to self-treat a condition, data on atomoxetine exposure reports from the National Poison Database System (NPDS) provides evidence that some who diverted atomoxetine had the intention of abuse and may have resulted in overdose. In a study of 20,032 atomoxetine exposures reported to the NPDS between 2002 and 2010, 10,608 (85.8%) cases were unintentional, 1,079 (8.7%) were reported as suicide attempts, and 629 (5.1%) cases were reported as abuse. Of the 12,370 single agent exposures, 21 major adverse medical events were reported, including eight tachycardic episodes, nine seizures, six comas, and one ventricular dysrhythmia. The authors concluded that although atomoxetine is generally safe, toxic side effects can occur and the risk of toxicity may be increased when patients are using atomoxetine for non-therapeutic purposes without monitoring by a health care provider (Monte et al., 2013).

Smaller-scale reports have corroborated these findings. A study in Texas showed that 6% of the poison control center calls for pediatric ingestion of atomoxetine resulted in serious outcomes, with one seizure reported (Forrester, 2007). Additionally, an analysis of regional poison control centers found 40 patients who had overdosed on atomoxetine. All patients were treated with activated charcoal and/or observation for symptoms of tachycardia, hypertension, vomiting, and drowsiness (Spiller, 2005). An additional study reviewed the effects of seventeen atomoxetine overdoses, with doses ranging from 10-1200 mg, which resulted in tachycardia, emesis, and agitation, with all symptoms resolving in 30 hours (Lovecchio & Kashani, 2006). Some studies have questioned whether atomoxetine carries a seizure risk, thus this has been an area of focus in several studies investigating the adverse effects associated with atomoxetine overdose (Kashani & Ruha, 2007; Wernicke et al., 2007). A 17 year old female ingested almost 3000 mg of atomoxetine, which resulted in a tonic clonic seizure and mild widening of the QRS interval (Kashani & Ruha, 2007). A review of clinical trial data showed adverse effects rates of seizures between 0.1-0.2%, which was not significantly different from the placebo group, and thus concluded that atomoxetine does not seem to elevate the risk of seizures in children with ADHD (Wernicke et al., 2007). During a 2-year period when 2.233 million adult and pediatric patients were exposed to atomoxetine, 8 out of 100,000 patients reported seizures (Reed et al., 2016).

In addition to seizures, a risk of cardiac events has been reported in numerous studies; however, mild increases in heart rate and blood pressure have been the most common (Reed et al., 2016; Henneson et al., 2017; Yu, 2022). A review of atomoxetine clinical trials found that of 8,417 patients, most pediatric patients experienced modest increases in heart rate (<20 bpm and <15 to 20 mmHg) and blood pressure and 8–12% experienced large increases (≥20 bpm and ≥15 to 20 mmHg) (Reed et al., 2016). However, after two years, blood pressure normalized and few patients discontinued atomoxetine due to cardiovascular adverse effects. QT interval prolongation, which is a concern associated with seizure risk, was found to be uncommon, affecting only 1.4% of 711 children and adolescents after three or more years of atomoxetine treatment (Reed et al., 2016). Currently, the US label and the European Supplementary Protection Certificate (SPC) warn atomoxetine users of an increase in heart rate and blood pressure. However, additional studies conducted on both children and adults have concluded that heart rate and blood pressure normalize with long-term use of atomoxetine (Yu, 2022; Reed et al., 2016). Therefore, previous findings strongly suggest that the risk of cardiac events for atomoxetine use is minimal, but patients’ heart rate and blood pressure should be monitored and a cardiovascular risk assessment performed before prescribing (Hennissen et al., 2017; Dadashova & Silverstone, 2012).

## Limitations

The findings in this study should be interpreted cautiously due to the relatively infrequent number of atomoxetine submissions, particulary for the generic formulation. An additional limitation is that many submissions were limited to Michigan and Pennsylvania, while thirteen states, primarily in the Midwest, had zero submissions. Additionally, StreetRx collects user-submitted data which may or may not be fully representative of the entire black market.

## Conclusions

This analysis identified 113 atomoxetine submissions with a street price of $1.35/mg. Additional pharmacodynamic studies on atomoxetine’s mechnism of action could help to explain its effects and thus elucidate why it might be desirable for cognitive enhancement or recreational use. Future qualitative studies should investigate the reasons why individuals are purchasing diverted atomoxetine on the black market (i.e. for recreational vs. nootropic vs. other clinical uses). If it is for therapeutic use (e.g. purchasing the drug on the black market to self-treat ADHD) or recreational use is identified, public health strategies could respond by either expanding legitimate atomoxetine access or limiting diversion.

## Data Availability

The data in this manuscript may be available. upon request to StreetRx.com.

https://streetrx.com/

