## Supplemental Figure 1 for "No stimulant? No Problem: Analyzing Black Market Sales of Atomoxetine on StreetRx"

**Supplemental Figure 1**. Atomoxetine average price per milligram superimposed on a violin plot as reported to StreetRx (Median = $0.05, Min = $0.01, Max = $20.00).

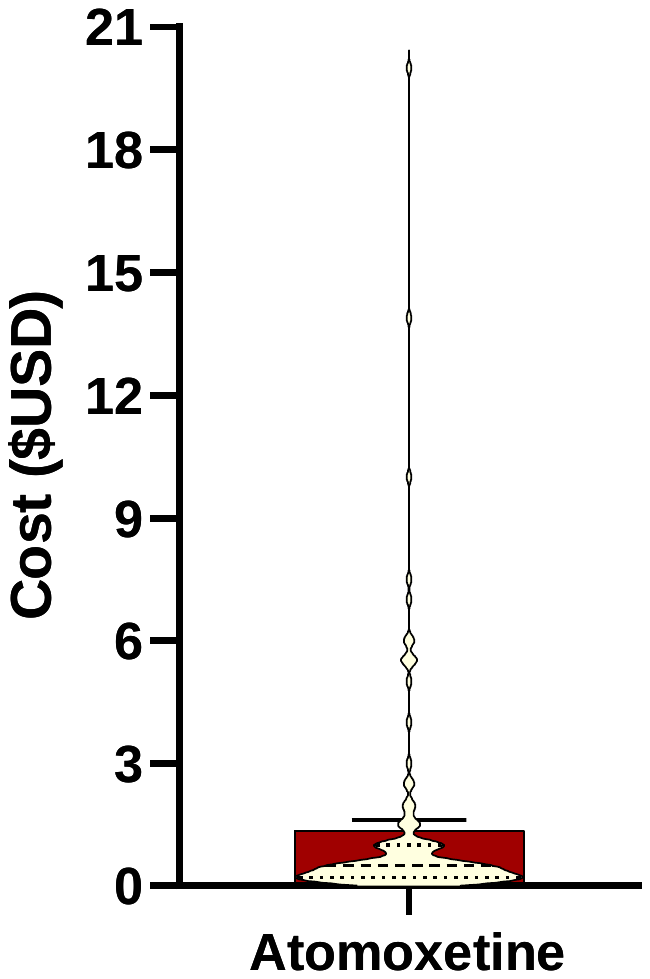
